# *Plasmodium falciparum* genomic surveillance reveals spatial and temporal trends, association of genetic and physical distance, and household clustering

**DOI:** 10.1101/2020.12.24.20248574

**Authors:** Mouhamad Sy, Awa Deme, Joshua L. Warren, Rachel F. Daniels, Baba Dieye, Pape Ibrahima Ndiaye, Younous Diedhiou, Amadou Moctar Mbaye, Sarah K. Volkman, Daniel L. Hartl, Dyann F. Wirth, Daouda Ndiaye, Amy K. Bei

## Abstract

Molecular epidemiology using genomic data can help identify relationships between malaria parasite population structure, malaria transmission intensity, and ultimately help generate actionable data to assess the effectiveness of malaria control strategies. Genomic data, coupled with geographic information systems data, can further identify clusters or hotspots of malaria transmission, parasite genetic and spatial connectivity, and parasite movement by human or mosquito mobility over time and space. In this study, we performed longitudinal genomic surveillance in a cohort of 70 participants over four years from different neighborhoods and households in Thiès, Senegal—a region of exceptionally low malaria transmission (entomological inoculation rate (EIR) less than 1). Genetic identity (identity by state) was established using a 24 single nucleotide polymorphism molecular barcode and a multivariable linear regression model was used to establish genetic and spatial relationships. Our results show clustering of genetically similar parasites within households and a decline in genetic similarity of parasites with increasing distance. One household showed extremely high diversity and warrants further investigation as to the source of these diverse genetic types. This study illustrates the utility of genomic data with traditional epidemiological approaches for surveillance and detection of trends and patterns in malaria transmission not only by neighborhood but also by household. This approach can be implemented regionally and countrywide to strengthen and support malaria control and elimination efforts.

## Introduction

As malaria transmission declines, and as population level immunity wanes, spatiotemporal variation in malaria incidence is likely to become more pronounced^1–4^. This heterogeneity is evidenced by hotspots of malaria transmission which can sustain and amplify the malaria transmission chain^5, 6^. Targeting and eliminating malaria hotspots can play an important role in malaria elimination^3, 6^. Different studies using conventional malaria epidemiologic tools such as malaria incidence and prevalence rates, malaria morbidity and mortality rates, and entomological inoculation rates (EIR) have identified hotspots of malaria transmission; however, some of these metrics are better suited for more highly endemic regions^7–9^ and may become less useful as malaria transmission declines. As some regions strive for malaria elimination, along with the decline of the disease incidence some of these conventional malaria metrics may become less informative^10^.

Genomic data from sensitive molecular tools are capable of detecting low level parasitemia and of providing additional information on parasite genetic population structure to measure the dynamic changes in malaria transmission^10, 11^. Genomic epidemiology has been used detect associations between malaria parasite genetic diversity, dynamic changes in transmission intensity, and malaria programmatic impact^12–15^. We have previously validated a 24-SNP molecular barcode for monitoring changes in transmission intensity as well as for tracking specific parasite types in the population^16^. More specifically, genomic tools can reveal whether specific genotypes dominate hotspots from focal local transmission of individual strains, or whether the malaria transmission landscape is characterized by increased genetic diversity with significant potential for outcrossing resulting from sustained transmission or importation of multiple genotypes^10, 17^. Genomic data can be coupled with mapping data, such as GIS, for visualizing spatial epidemiology. These combined data types can be informative for evaluating control measures. For example, GIS and epidemiologic data were used for mapping clusters of malaria transmission in Gambia, Mali and Senegal and used to guide optimal malaria control intervention^18^. GIS information can also give important information at the household level for studies in micro-epidemiology and ecology^19^. This combined approach was used in Papua Indonesia by coupling genetic information of *P. falciparum* and *P. vivax* from microsatellite data and household level GIS information to study malaria micro-epidemiology^20^. In Senegal, few studies have used genomic data to understand *Plasmodium* parasite genetic diversity and spatiotemporal dynamics. Here, we seek to bridge this gap by using genomic epidemiology and ecology at the city, neighborhood, and household levels.

The aim of this present study was to apply the 24-SNP molecular barcode in a longitudinal cohort enrolled between 2014 and 2017 and followed for 2 years after enrollment to understand *P. falciparum* parasite population structure in Thiès, Senegal. We determined the spatio-temporal parasite haplotype distribution, and the association between physical distance and parasite genetic distance and the interconnectivity between parasites using multivariable linear regression models. These analyses helped generate hypotheses on possible reasons for transmission hotspots. The overall goal of this study is to help inform malaria control by integrating genomic data into decision making.

## Results

### Patient demographics and characteristics of the cohort

A total of 70 participants were enrolled following informed consent spanning 4 years (2014 = 2, 2015 = 32, 2016 = 23 and 2017 = 13) and followed for two years post-enrollment. Patients were recruited through passive case detection upon presenting at the Service de Lutte Anti Parasitaire (SLAP) clinic with malaria-like symptoms and testing positive by malaria rapid diagnostic test (Pfhrp2 antigen RDT) and microscopy for P. falciparum monogenomic infection. Patients were residents of 6 different neighborhoods in Thiès (Cité Senghor, Diakhao, Escale, Nguinth, Thialy and Takhikao) and 10 different houses (BD, BS, DL, DMS, GB, MS, OB, OD, SD and SN). The majority of the participants were from Diakhao 51/70 (72.8%), and the majority of shared household participants were in household MS; 37/70 (52.8%) (Table 1). While enrollment was open to all genders and ages, in this study all participants were male aged from 5 to 16 years. When evaluating the gender bias after enrollment, many were living in “daaras” – the equivalent of religious boarding schools. The mean age was 10.91 years, and 25% (18/70) of the participants were under 10 years. The mean parasitemias were 0.77%, with a minimum of 0.03% and a maximum of 4.89% (Table 1).

**Table 1.** Demographic Characteristics of the Study Population. Patient demographics for the n=70 participants. Sex, Age (in years), Parasitemia (calculated from thin smears), and breakdown of participants by neighborhood are shown.

| Characteristic | Number (%) |
| --- | --- |
| **Sex, Number (%)** |  |
| Male | 70 (100) |
| Female | 0 |
| Ratio (M/F) | – |
| **Age, years** |  |
| Mean | 10.91 |
| Range | (5 – 16) |
| **Parasitemia, (%)** |  |
| Mean | 0.77% |
| Range | (0.03% – 4.89%) |
| **Neighborhood, No. (%)** |  |
| Cite Senghor | 1 (1%) |
| Diakhao | 47 (67%) |
| Escale | 1 (1%) |
| Takhikao | 5 (7%) |
| Thialy | 5 (7%) |
| Nguinth | 11 (15%) |

### Limited genetic diversity and high frequency of monogenomic infection

Among the 70 participants, there was a total of 74 distinct infections. All participants were parasitemic at day 0, and 4 participants were infected with a subsequent re-infection over the course of 2 years of follow-up (8 visits, 4 each year). Overall, monogenetic infection was predominant 90.5% (67/74) and polygenomic infections were rare, 9.4% (7/74). This finding is predicted for the region^14^. The majority of polygenomic infections were observed in 2015 (5/7). More than 41.8% (31/74) of parasite genetic types were shared within participants overall (2014 = 0/2, 2015 = 11/32, 2016 = 12/27 and 2017 = 8/13). Twenty-seven haplotypes previously described in Thiès were detected in 56 isolates (56/74); nine unique genotypes were observed in this cohort and detected in eleven participants (11/70). Among the 27 haplotypes we found 8 clusters (shared parasite haplotypes between at least two participants); these were haplotypes 25, 83, 759, 796, 804, 846, 873, and 1001. Among the 19 unique parasites, 2 clusters (U2 and U4) in a total of 4 infections were found (Figure 1). A heatmap reveals parasite clustering by barcode similarity (Figure 2).

**Figure 1.**
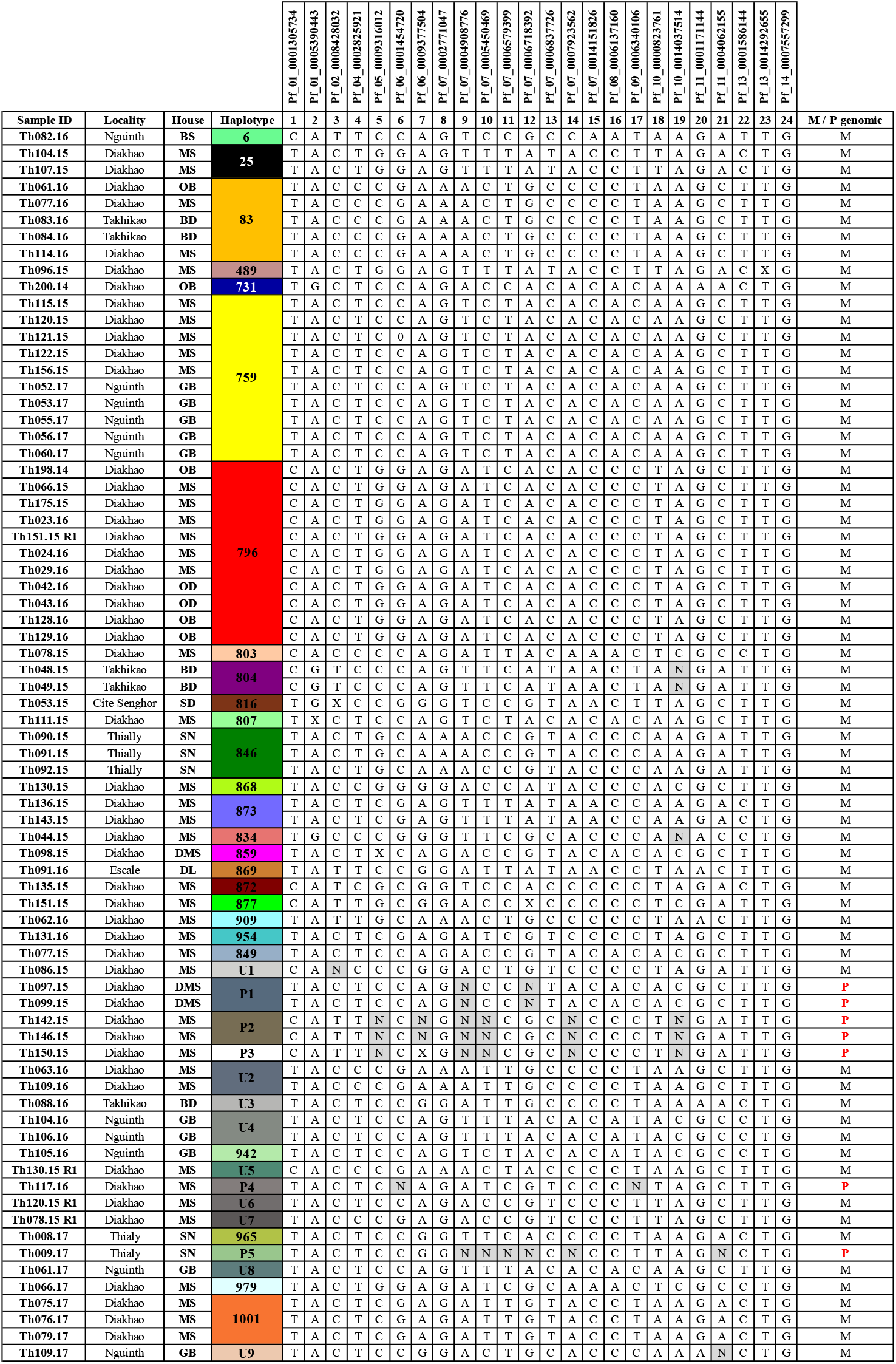
Barcodes of Plasmodium falciparum isolates Identified in the Study Cohort. Barcoded parasites from the n=70 participants in the longitudinal cohort followed for 24 months. Patient ID (human) is shown along with date of sampling, Age, Neighborhood, Household (arbitrary alphabetical code), and barcode haplotype. SNP positions are indicated for the 24-SNP barcode. “N” indicates a position with two peaks – corresponding to both biallelic SNPs (mixed genotype), and an “X” indicates a reproducibly negative call. M/P genomic indicates monogenomic (M) or polygenomic (P) infections.

**Figure 2.**
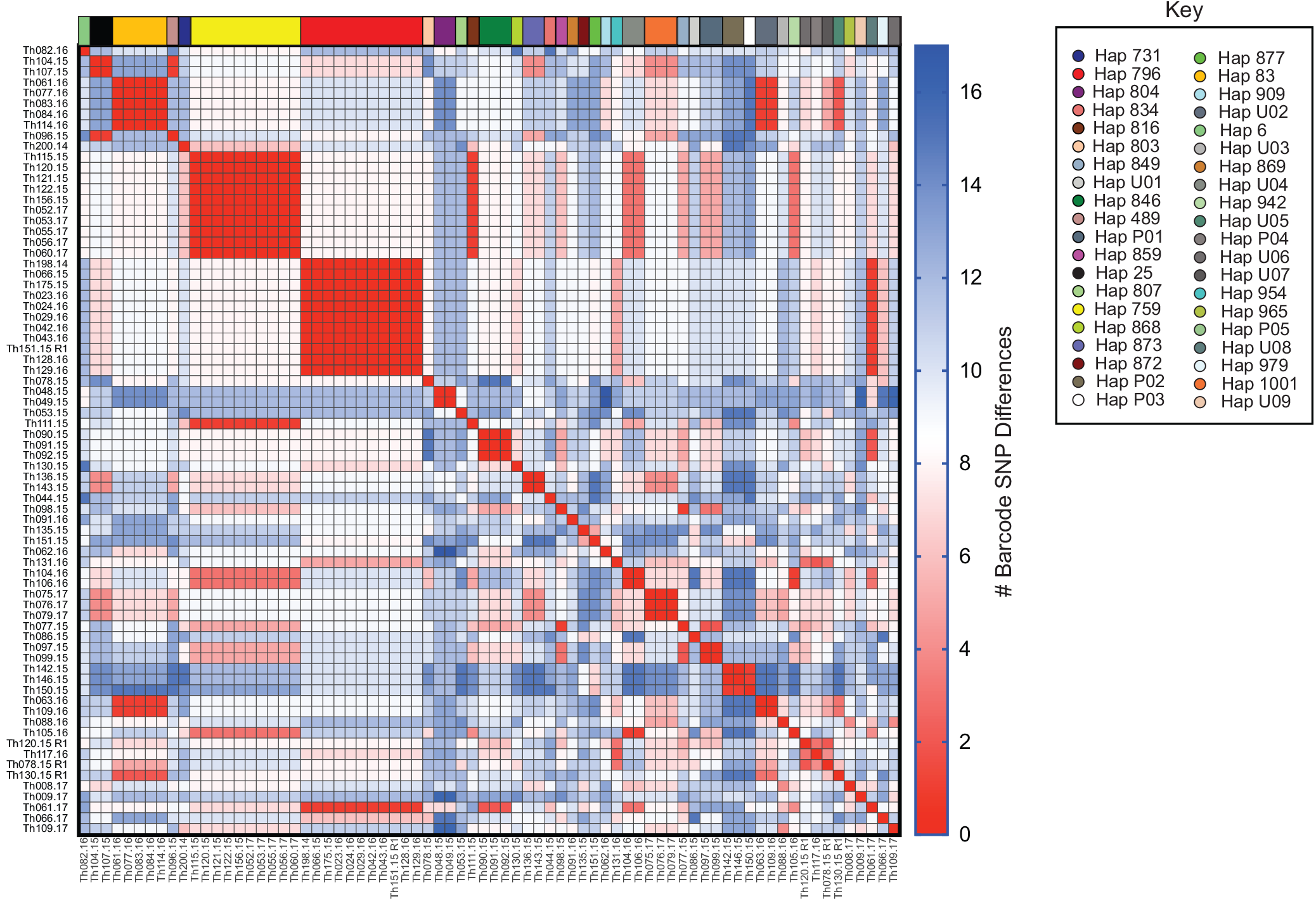
Heatmap illustrating parasite genetic barcode similarity. Sample IDs are listed on the vertical axis and barcodes are clustered by genetic similarity. Barcode haplotypes are color coded at the top of the heatmap and in the Key. Heatmap scale (right) shows the number of barcode SNP differences and range from identical (fewest differences; red) to greatest differences (blue). The maximal number of differences possible is 24.

### Genomic haplotypes are shared within years and persist across malaria transmission seasons

Genetic types were shared within a transmission season but also across transmission seasons (lineage persistence). The dominant haplotypes that persisted for multiple seasons were haplotypes 759 and 796. Haplotype 759 (n = 10) was detected in 2015 in Diakhao in the MS household and in 2017 in Nguinth in the GB household. Haplotype 796 (n = 11) was observed in 2014 (n = 1), 2015 (both in MS) and 2016 (n = 8) in the neighborhood of Diakhao in three different households (MS, OB and OD). Haplotype 83 was found in 2016 in two different neighborhoods (Diakhao and Takhikao) in three different households (MS, OB and BD) (Figure 3).

**Figure 3.**
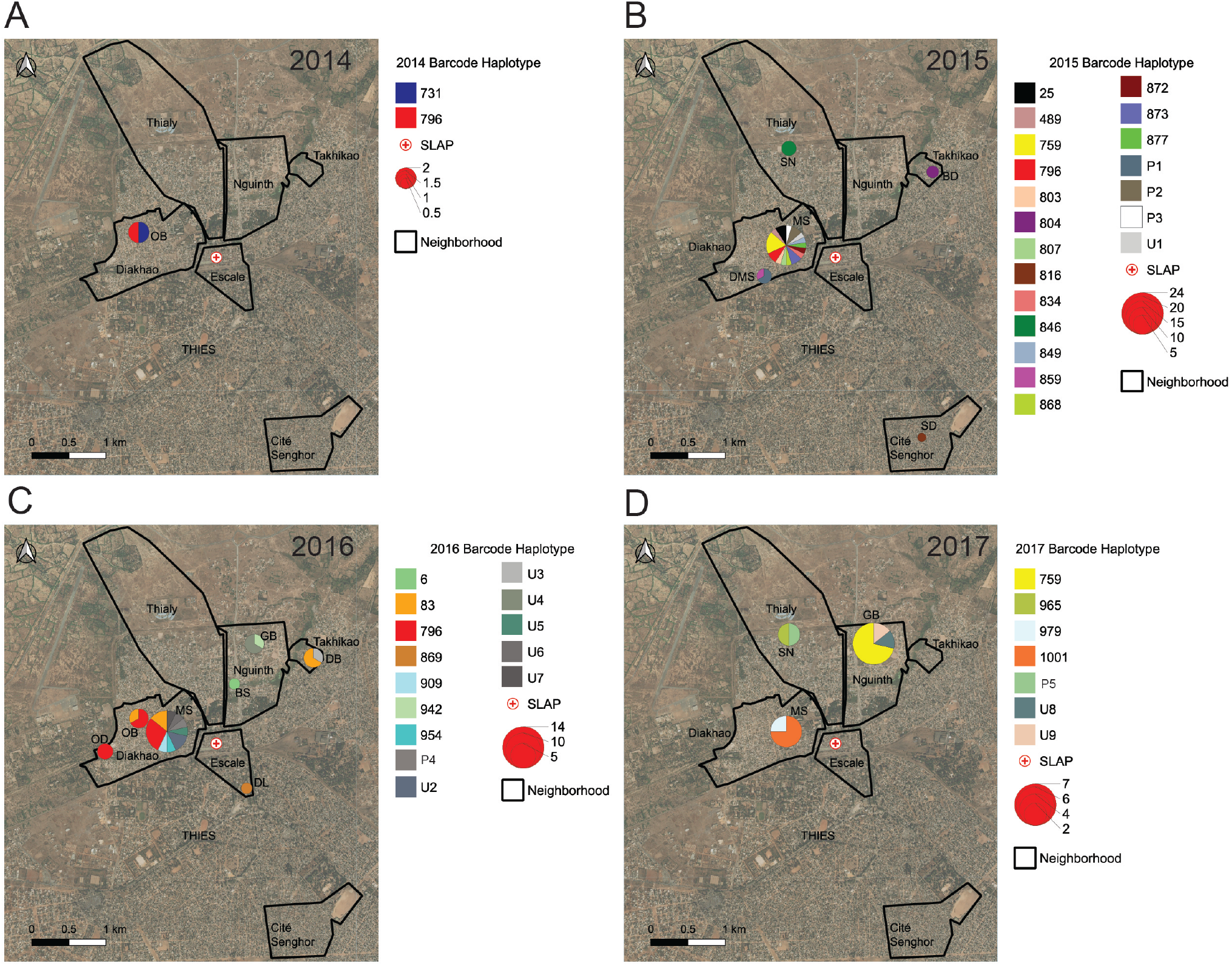
Hotspots of malaria transmission by neighborhood and household. Map of sample locations by household (BD, BS, DL, DMS, GB, MS, OB, OD, SD and SN), and neighborhood (Cité Senghor, Diakhao, Escale, Nguinth, Thialy and Takhikao) from 2014 to 2017. Legend indicates and the parasite barcode haplotype ID and is color-coded by haplotype. The size of the circle is proportional to the number of samples in the haplotype, as indicated in the scale. Solid lines indicate delineation of the neighborhoods.

### Genomic signatures among initial and re-infection

During the patient follow-up, 4 participants recruited in 2015 (Th078.15, Th120.15, Th130.15 and Th151.15) were re-infected in the following year during the peak of the malaria transmission period. Unscheduled visits in which study subjects had symptomatic infections are designated as “re-infections” or R1 (Th078.15.R1, Th120.15.R1, Th130.15.R1 and Th151.15.R1). Re-infected participants were all from the same household (MS). Parasite genotypes from the first infection were genetically distinct from the re-infected parasites (Figure 4), and three of them (U5, U6 and U7) represented genotypes which had not been previously described, either in Thiès or in multiple regions of Senegal from 2006-present^14, 16, 21^.

**Figure 4.**
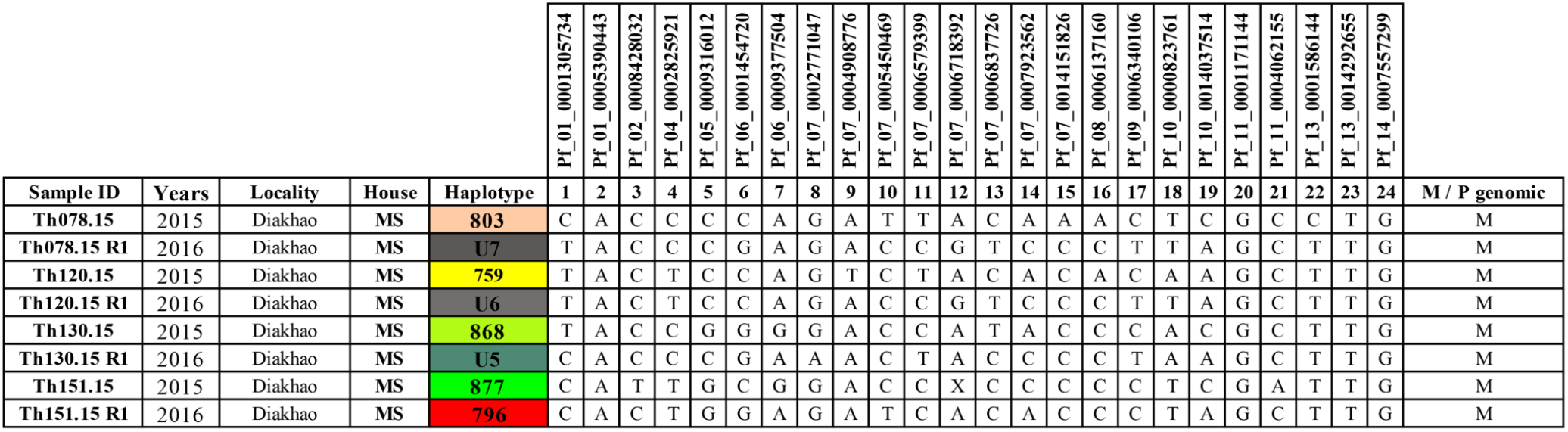
Barcodes of initial and re-infected participant samples. Patient ID (human) is shown along with date of sampling, Age, Neighborhood, Household (arbitrary alphabetical code), and barcode haplotype. SNP positions are indicated for the 24-SNP barcode. “N” indicates a position with two peaks – corresponding to both biallelic SNPs (mixed genotype), and an “X” indicates a reproducibly negative call. M/P genomic indicates monogenomic (M) or polygenomic (P) infections.

### Effect of spatial distance on parasite genomic similarity

We calculated the genetic similarity between each unique pair of patients as well as the geographic distance between their respective households to determine whether increasing physical distance between households is associated with greater genetic difference. When analyzed by year (to normalize for the circulating genetic variants present within a season), we observed a significant positive association between physical and genetic distance in 2015 and 2016 (Table 2); as physical distance increased between two households, so did the genetic distance of the patients in those households. When data were analyzed within a year (to account for differences in the genetic types most prevalent between years), we observed the same trend. Further, we observed that for all households except one (MS), living in the same household was more likely to result in participants being infected with similar genomes, although these findings were not statistically significant for all households (Table 2). In all years, there were signs of housing cluster effects, with genetic distance decreasing significantly for participants located in certain houses together.

**Table 2.** Relationship between physical distance and genetic distance. The relationship between physical distance (measured in kilometers (km) between two GPS coordinates) and genetic distance (measured as identity by state, the number of barcode differences in the 24-SNP molecular barcode) are shown overall, as well as by household. Parameter estimates are shown, with standard error (SE). Positive values indicate a positive association (greater genetic distance) and negative values indicate a negative association (less genetic distance, parasites are more similar). P-values from multivariable linear regression indicate whether the parameter is significantly different from zero, and are calculated using a significance value of alpha = 0.05.

| **Effect (2015)** | **Estimate (SE)** | **P-value** |
| --- | --- | --- |
| Distance (km) | 0.40 (0.19) | 0.04 |
| House 1: BD | -0.54 (2.97) | 0.86 |
| House 2: DMS | -0.54 (1.73) | 0.76 |
| House 3: MS | -0.00 (0.36) | 0.99 |
| House 4: SN | -4.87 (1.73) | 0.01 |
| **Effect (2016)** | **Estimate (SE)** | **P-value** |
| Distance (km) | 1.00 (0.50) | 0.04 |
| House 1: BD | -6.33 (2.28) | 0.01 |
| House 2: GB | -1.66 (2.28) | 0.47 |
| House 3: MS | 0.56 (0.85) | 0.51 |
| House 4: OB | -0.33 (2.28) | 0.89 |
| House 5: OD | -6.99 (3.85) | 0.07 |
| **Effect (2017)** | **Estimate (SE)** | **P-value** |
| Distance (km) | -2.12 (3.77) | 0.58 |
| House 1: BD | -6.90 (2.81) | 0.02 |
| House 2: GB | -6.11 (2.95) | 0.04 |
| House 3: MS | -3.61 (3.77) | 0.34 |

## Discussion

As malaria control progresses towards elimination, genomic data has proven to be essential in assessing control and surveillance^22^. In this study we combine parasite genetic diversity indices, individual GPS information at the household and neighborhood level, and multivariable linear regression models to understand parasite diversity and connectivity over time and space in Thiès, Senegal. The main findings of this study were household clustering of genetic types, association with genetic distance and physical distance, as well as parasite sharing between participants from either the same household or different households which were geographically proximal. Interestingly, in past analyses from cross-sectional studies in Thiès, this spatial clustering of identical genotypes was not observed^23^.

Overall, the low level of *Plasmodium* parasite genetic diversity and the high frequency of monogenomic infection observed over years are generalizable and consistent with previous observations in the longitudinal cross-sectional sampling over time in Thiès^14, 23^ and Dielmo and Ndiop in Senegal^16^. In these localities 24-SNP molecular barcoding revels a predominance of monogenic infection and a significant percentage of shared genomic haplotypes in the population. These observations have been hypothesized to be the result of a significant reduction in malaria transmission due to the efficiency of malaria interventions post-2008. Malaria parasites are known to evolve quickly, in part due to sexual recombination in mosquitos, which plays an important role in parasite population structure and complexity of infection^17^. As expected, in this low transmission setting, we see decreased evidence of outcrossing and increased selfing of identical parasites relative to populations with higher transmission intensity and a higher complexity of infection^14, 24^.

Consistent with previous studies in Thiès^14, 23^, we also observed the existence of haplotypes persisting over several years. Our study adds the additional GIS data which permitted us to monitor the genotype frequencies in different households and neighborhoods within the same year and across seasons. We also have the example of haplotype 796 observed in the same neighborhood (Diakhao) in 3 successive years (2014 - 2016) in three different households (OB, OD, and MS). When observing identical genotypes in households, there are two possibilities: 1) continued local transmission of a single parasite genotype, or 2) a single infected mosquito biting multiple infected individuals within the household. The temporal nature of the infections can help distinguish which hypothesis is more likely. This study has also demonstrated that simultaneous transmission of the same genotype is frequent in Thiès^25^.

We observed household clustering and genetic differences between parasites to increase with distance between individuals. During this study, a particular household (MS) served as an example of a malaria hotspot of transmission at the household level, both in number of cases as well as genetic diversity of the parasites. Having such different parasites in the same household could be the result of a hotspot of local intense transmission^17^, coupled with genetic recombination (outcrossing) within the Anopheles mosquito^26^, and the subsequent transmission of new genetic combinations^17^. An alternative could be the importation of diverse genotypes due to human or mosquito mobility^21, 27–30^. A similar study of malaria incidence and prevalence has demonstrated the existence of malaria transmission hotspots at the village level in Senegal^7, 8^. In such villages, human density, human behavior, lax malaria bed-net use, substandard housing construction, and a favorable ecological environment for mosquito proliferation (presence of mosquito breeding sites) have all been identified as risk factors for a household to be in a hotspot^31^. The added value of our approach is being able to identify hotspots of transmission in terms of cases, but also to determine the genotypic nature of these hotspots – adding further to implications for control measures. If hotspots are populated by similar genotypes, it is more likely that local transmission is occurring. If multiple diverse genotypes are present, the hotspot could serve as a hub of human or mosquito imported infections. Identification of the transmission clusters at the household level will play an important role for interrupting malaria transmission chains^5, 32^. In our study, the prevention and control interventions needed to combat malaria in a household with a homogenous genetic haplotype may be very different from a household (such as MS) where many genetically diverse parasites are prevalent. This study provides new support of how molecular tools can help identify malaria hotspots.

Because *P. falciparum* is a sexually recombining organism, precise mapping of phylogeny and transmission chains is not possible; however, the 24-SNP barcode has been shown to be a proxy for whole-genome that allows resolution especially of highly similar parasite types^14^. While the 24-SNP barcode does not provide as complete information about genetic relatedness (identity by descent) as whole-genome sequencing or large SNP arrays^33^, it has been estimated that the 24-SNP barcode can confidently detect parasites that share greater than 70% genome similarity (identity by state)^14^.

While the pairwise genetic distance in the 24-SNP barcode is not linearly associated with whole-genome genetic distance, our finding of significant associations with physical distance is even more noteworthy. Our statistical model demonstrated that genetic variation between parasite pairs increases with physical distance. Here we used the number of SNP differences between paired individuals as genetic distance, or identity by state. Studies in The Gambia and Kenya have demonstrated that variation between parasite genotypes increases with geographical distance^34, 35^. Such findings will help in understanding how the parasite population is structured in Thiès and the connectivity between parasites, despite some studies in Thiès having suggested a mixed parasite population with no hidden population structure^25^. In this study, sampling biases (number of limited samples) may not reflect the overall parasite population that is captured by passive case detection, and notably, we found no asymptomatic infections in any of the follow-up time points in the cohort.

Many of the enrolled participants in our study live in “*daara*”, religious boarding schools where “*talibe*” (student followers) live together in large numbers. Our cohort was completely male, although enrollment was open and encouraged for both male and females. These limitations may affect the generalizability of the study beyond these populations. Nonetheless, our methods provide important information in the micro-epidemiology of parasite population structure in space and time in Thiès. The study also provides evidence of the feasibility and power of including genomic analyses in making public health decisions.

In this study malaria spatial-temporal clustering at the household and neighborhood level were observed along with increasing genetic distance between parasites as a function of physical distance. The longitudinal study shows the importance of applying molecular surveillance along with spatial and temporal modeling to detect hotspots of malaria transmission. This approach can be applied at a small scale as well as at a regional scale and countrywide for detecting and targeting malaria foci to further assess initiatives of malaria control and intervention.

## Methods

### Ethics Statement

Ethical approval for this study was granted by the National Ethics Committee of the Ministry of Health in Senegal (Protocol SEN 14/49), the Institutional Review Board of the Harvard T.H. Chan School of Public Health (IRB 14-2830), and the Human Investigation Committee of Yale University (Protocol 2000023287). All samples were collected with informed consent and in accordance with all ethical requirements of the National Ethics Committee of Senegal, Institutional Review Board of the Harvard T.H. Chan School of Public Health, and the Harvard, and the Human Investigation Committee of Yale University.

### Inclusion and Exclusion Criteria

Samples were collected from patients greater than 6 months of age with uncomplicated *P. falciparum* malaria confirmed by a peripheral blood smear and rapid diagnostic test (RDT) by the local health officer of the health clinic of Thiès (SLAP). Patients were asked to return to the health post, regardless of health status, at days 1, 2, 3 and after 2 weeks, 4 weeks, 3 months, 6 months, 12 months, 18 months, and 24 months. Patients were also asked to return for an unscheduled visit if they experienced malaria-like symptoms. At the visit on days 1-3, the patient was monitored for the clearance of parasitemia by finger prick and a microscopy slide and an RDT was evaluated. At scheduled follow-up visits at 2 weeks, 4 weeks (1 month), 3 months, 6 months, 12 months, 18 months, and 24 months, 5 mLs of blood was drawn for plasma and PBMCs. On Day 0 and at unscheduled visits where a patient was confirmed to be positive with *P. falciparum*, blood was also cryopreserved, and parasite DNA was extracted from whole blood with the QIAamp DNA blood mini kit (Qiagen Inc., Valencia, CA, USA).

### Molecular Barcoding genotyping

24 SNP molecular barcodes were identified using a previously described assay^36^. Barcode assays were run on the LightCycler 96 Roche system. SNPs were amplified as follows; 2.0 *µ*L of Lightscanner Master Mix (BioFire Defense), 2.5 *µ*L of a 1:100 dilution DNA template, and 0.5 *µ*L of primers and probes. Genomic DNA from cultured *P. falciparum* strains (3D7, Dd2, 7G8, Tm90) was used for assay validation and as genotyping controls for all reaction plates. Molecular barcode assays 10, 11, 13, 21, and 24 were performed optimally under asymmetric forward to reverse primer ratios of 5:1; all other assays required a 1:5 primer asymmetry. Amplification conditions were 95^°^C denaturation for 2 minutes, 50 cycles of 94^°^C for 5 seconds and 66^°^C for 30 seconds, plus a pre-melt cycle of 5 seconds each at 95^°^C and 37^°^C. Two or more N’s among the 24 SNPs assayed was taken to indicate that more than one *P. falciparum* genomes was present (a polygenomic infection). Ambiguous calls and calls with “X” were repeated 3 times in independent experiments before validation^36^.

### GIS analysis and statistical modeling

GPS coordinates of participants’ households (while not revealing individual participant addresses or identifiable locations) and neighborhoods were used to make different maps with QGIS 3 (http://www.qgis.osgeo.org). We used multivariable linear regression models to determine if the genetic similarity between pairs of participants is related to the geographic distance that separates them. The number of 24-SNP barcode differences between each unique pair of participants was used to describe their genetic similarity. We used two metrics to describe spatial proximity in the analysis. First, for each unique pair of participants we determined whether the individuals were located in the same house and if so, noted which house it was. Next, we calculated the geographic distance between the house centroids for each pair. In this way, we explore the impact of geographic distance on genetic similarity in two ways; are people clustered in the same house more genetically similar and are individuals in houses that are closer together geographically more genetically similar.

We then model genetic distance between each pair of participants as a function of the spatial distance between their houses and a clustering indicator for the specific house, where each house has its own specific regression parameter. The model is given as:

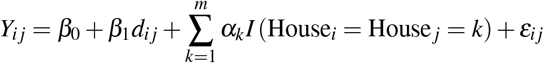

where *Y*_*ij*_ is the genetic distance between participants *i* and *j, d*_*ij*_ is the geographic distance between the house centroids of participants *i* and *j, m* is the total number of unique houses in the analyzed dataset, *I*(.) is an indicator function that is equal to one if the input statement is true and is equal to zero otherwise, and *ε*_*ij*_ ∼ N (0, *σ*^2^). This model relaxes the assumption that clustering in any house has the same impact on genetic similarity and allows for the possibility that this effect changes across the different houses (i.e., *α*_*k*_). The parameter *β*_1_ describes the association between genetic and geographic distance between houses. We fit this model to each individual year of data separately and present inference for the model parameters^37, 38^.

## Data Availability

Data associated with this manuscript can be found at: https://doi.org/10.5061/dryad.wh70rxwmk

## Acknowledgements

We would like to express great gratitude to the population of Thiès, Senegal and to the heath workers of the SLAP clinic for their invaluable collaboration and contribution to this work. We sincerely appreciate the helpful discussions and useful inputs made by Daniel Neafsey to this study. We would like to thank Sidiya Mbodj and Fatoumata Dabo for assistance with geolocalization. We sincerely thank all colleagues who contributed to this work. Funding for the study was provided by the NIH (K01 TW010496) to Amy K. Bei, and by the Bill and Melinda Gates Foundation (OPP1053604) to Dyann F. Wirth.

## Data Accessibility

Data associated with this manuscript can be found at: https://doi.org/10.5061/dryad.wh70rxwmk.

## Author contributions statement

A.K.B, D.N. and D.F.W conceived and designed the study. B.D, P.I.N, Y.D. and A.M.M. collected and processed samples and managed patient databases. M.S., A.B.D, and A.K.B performed molecular barcoding and genomic analysis and interpretation. M.S and A.K.B performed mapping and data visualization. J.W. performed data modeling and analysis. M.S., J.W., S.K.V, D.L.H, A.K.B. revised this manuscript. All authors read and approved the final manuscript.

## References

1. World malaria report 2020: 20 years of global progress and challenges. Report, World Health Organization (2020).

2. Fowkes, F. J., Boeuf, P. & Beeson, J. G. Immunity to malaria in an era of declining malaria transmission. Parasitology 143, 139–53, DOI: 10.1017/S0031182015001249 (2016).

3. Nkumama, I. N., O’Meara, W. P. & Osier, F. H. A. Changes in Malaria Epidemiology in Africa and New Challenges for Elimination. Trends Parasitol 33, 128–140, DOI: 10.1016/j.pt.2016.11.006 (2017).

4. Cotter, C. et al. The changing epidemiology of malaria elimination: new strategies for new challenges. Lancet 382, 900–11, DOI: 10.1016/S0140-6736(13)60310-4 (2013).

5. Stresman, G., Bousema, T. & Cook, J. Malaria Hotspots: Is There Epidemiological Evidence for Fine-Scale Spatial Targeting of Interventions? Trends Parasitol 35, 822–834, DOI: 10.1016/j.pt.2019.07.013 (2019).

6. Bousema, T. et al. Hitting hotspots: spatial targeting of malaria for control and elimination. PLoS Med 9, e1001165, DOI: 10.1371/journal.pmed.1001165 (2012).

7. Dieng, S. et al. Spatio-temporal variation of malaria hotspots in Central Senegal, 2008-2012. BMC Infect Dis 20, 424, DOI: 10.1186/s12879-020-05145-w (2020).

8. Ndiath, M. et al. Identifying malaria hotspots in Keur Soce health and demographic surveillance site in context of low transmission. Malar J 13, 453, DOI: 10.1186/1475-2875-13-453 (2014).

9. Tusting, L. S., Bousema, T., Smith, D. L. & Drakeley, C. Measuring changes in Plasmodium falciparum transmission: precision, accuracy and costs of metrics. Adv Parasitol 84, 151–208, DOI: 10.1016/B978-0-12-800099-1.00003-X (2014).

10. Neafsey, D. E. & Volkman, S. K. Malaria Genomics in the Era of Eradication. Cold Spring Harb Perspect Med 7, DOI: 10.1101/cshperspect.a025544 (2017).

11. Lucchi, N. W., Oberstaller, J., Kissinger, J. C. & Udhayakumar, V. Malaria diagnostics and surveillance in the post-genomic era. Public Heal. Genomics 16, 37–43, DOI: 10.1159/000345607 (2013).

12. Anderson, T. J. et al. Microsatellite markers reveal a spectrum of population structures in the malaria parasite Plasmodium falciparum. Mol Biol Evol 17, 1467–82, DOI: 10.1093/oxfordjournals.molbev.a026247 (2000).

13. Mwingira, F. et al. Plasmodium falciparum msp1, msp2 and glurp allele frequency and diversity in sub-Saharan Africa. Malar J 10, 79, DOI: 10.1186/1475-2875-10-79 (2011).

14. Daniels, R. F. et al. Modeling malaria genomics reveals transmission decline and rebound in Senegal. Proc Natl Acad Sci U S A 112, 7067–72, DOI: 10.1073/pnas.1505691112 (2015).

15. Wesolowski, A. et al. Mapping malaria by combining parasite genomic and epidemiologic data. BMC Med 16, 190, DOI: 10.1186/s12916-018-1181-9 (2018).

16. Bei, A. K. et al. Dramatic Changes in Malaria Population Genetic Complexity in Dielmo and Ndiop, Senegal, Revealed Using Genomic Surveillance. J Infect Dis 217, 622–627, DOI: 10.1093/infdis/jix580 (2018).

17. Koepfli, C. & Mueller, I. Malaria Epidemiology at the Clone Level. Trends Parasitol 33, 974–985, DOI: 10.1016/j.pt.2017.08.013 (2017).

18. Shaffer, J. G. et al. Clustering of asymptomatic Plasmodium falciparum infection and the effectiveness of targeted malaria control measures. Malar J 19, 33, DOI: 10.1186/s12936-019-3063-9 (2020).

19. Garcia, G. A. et al. Mapping and enumerating houses and households to support malaria control interventions on Bioko Island. Malar J 18, 283, DOI: 10.1186/s12936-019-2920-x (2019).

20. Pava, Z. et al. Genetic micro-epidemiology of malaria in Papua Indonesia: Extensive P. vivax diversity and a distinct subpopulation of asymptomatic P. falciparum infections. PLoS One 12, e0177445, DOI: 10.1371/journal.pone.0177445 (2017).

21. Daniels, R. F. et al. Genetic evidence for imported malaria and local transmission in Richard Toll, Senegal. Malar J 19, 276, DOI: 10.1186/s12936-020-03346-x (2020).

22. Nsanzabana, C. Strengthening Surveillance Systems for Malaria Elimination by Integrating Molecular and Genomic Data. Trop Med Infect Dis 4, DOI: 10.3390/tropicalmed4040139 (2019).

23. Bei, A. K. et al. Plasmodium falciparum population genetic complexity influences transcriptional profile and immune recognition of highly related genotypic clusters. bioRxiv 2020.01.03.894220, DOI: 10.1101/2020.01.03.894220 (2020).

24. Nkhoma, S. C. et al. Population genetic correlates of declining transmission in a human pathogen. Mol Ecol 22, 273–85, DOI: 10.1111/mec.12099 (2013).

25. Wong, W. et al. Genetic relatedness analysis reveals the cotransmission of genetically related Plasmodium falciparum parasites in Thies, Senegal. Genome Med 9, 5, DOI: 10.1186/s13073-017-0398-0 (2017).

26. Miles, A. et al. Indels, structural variation, and recombination drive genomic diversity in Plasmodium falciparum. Genome Res 26, 1288–99, DOI: 10.1101/gr.203711.115 (2016).

27. Wesolowski, A., Eagle, N., Noor, A. M., Snow, R. W. & Buckee, C. O. Heterogeneous mobile phone ownership and usage patterns in Kenya. PLoS One 7, e35319, DOI: 10.1371/journal.pone.0035319 (2012).

28. Tessema, S. et al. Using parasite genetic and human mobility data to infer local and cross-border malaria connectivity in Southern Africa. Elife 8, DOI: 10.7554/eLife.43510 (2019).

29. Guerra, C. A. et al. Human mobility patterns and malaria importation on Bioko Island. Nat Commun 10, 2332, DOI: 10.1038/s41467-019-10339-1 (2019).

30. Roh, M. E. et al. High Genetic Diversity of Plasmodium falciparum in the Low-Transmission Setting of the Kingdom of Eswatini. J Infect Dis 220, 1346–1354, DOI: 10.1093/infdis/jiz305 (2019).

31. Bannister-Tyrrell, M. et al. Importance of household-level risk factors in explaining micro-epidemiology of asymptomatic malaria infections in Ratanakiri Province, Cambodia. Sci Rep 8, 11643, DOI: 10.1038/s41598-018-30193-3 (2018).

32. Bousema, T. et al. Identification of hot spots of malaria transmission for targeted malaria control. J Infect Dis 201, 1764–74, DOI: 10.1086/652456 (2010).

33. Taylor, A. R., Jacob, P. E., Neafsey, D. E. & Buckee, C. O. Estimating Relatedness Between Malaria Parasites. Genetics 212, 1337–1351, DOI: 10.1534/genetics.119.302120 (2019).

34. Amambua-Ngwa, A. et al. Long-distance transmission patterns modelled from SNP barcodes of Plasmodium falciparum infections in The Gambia. Sci Rep 9, 13515, DOI: 10.1038/s41598-019-49991-4 (2019).

35. Omedo, I. et al. Micro-epidemiological structuring of Plasmodium falciparum parasite populations in regions with varying transmission intensities in Africa. Wellcome Open Res 2, 10, DOI: 10.12688/wellcomeopenres.10784.2 (2017).

36. Daniels, R. et al. A general SNP-based molecular barcode for Plasmodium falciparum identification and tracking. Malar J 7, 223, DOI: 1475-2875-7-223[pii]10.1186/1475-2875-7-223 (2008).

37. Warren, J. L. et al. Investigating spillover of multidrug-resistant tuberculosis from a prison: a spatial and molecular epidemiological analysis. BMC Med 16, 122, DOI: 10.1186/s12916-018-1111-x (2018).

38. Yang, C. et al. Internal migration and transmission dynamics of tuberculosis in Shanghai, China: an epidemiological, spatial, genomic analysis. Lancet Infect Dis 18, 788–795, DOI: 10.1016/S1473-3099(18)30218-4 (2018).

